# Dysregulation of gut microbiota composition in individuals with personality disorders: A systemic review and meta-analysis

**DOI:** 10.1101/2023.07.19.23292891

**Authors:** Rangraze Imran, Shehla Khan

## Abstract

**Background:** Anxiety disorders are the most frequent mental comorbidity in people with functional GI difficulties, and abdominal discomfort is one of the most known physical signs of sadness. Successful top-down treatments using antidepressants and psychosocial therapies in the treatment of irritable bowel syndrome (IBS) further define personality illnesses as more than merely CNS disorders, but disorders with highly extensive systemic interconnections.

Therefore, we recently conducted a systematic review of the observational literature comparing the gut microbiota composition of persons with personality difficulties with healthy control.

**Methods:** This review was written according to the guidelines established by Preferred Reporting Items for Systematic Reviews and Meta-Analyses (PRISMA). Not a single rule was broken, yet a more thorough search strategy did provide more relevant results. Pubmed, Scopus, Embase, Web of Science, Ovid, Global Health, PsycINFO, etc. were searched thoroughly using the phrases “gut microbiota, psychological disorders, personality disorders, composition, major depressive disorder, bipolar disorders, schizophrenia, etc.”

**Results:** Researchers did discover widespread differences in the gut microbiota of patients and controls under each category of personality disorder. They also found that there are distinct bacterial taxa that had differing abundances in patients with these three psychiatric illnesses compared to healthy controls. They found a great deal of variation in study designs and reporting, such as in the inclusion and exclusion of study populations, sampling feces for study of gut microbiota; taking into account or adjusting for important factors known to impact gut microbiota composition; storing feces; processing feces; analyzing feces.

**Conclusion:** Our systematic review did find that psychological disorders appeared to exhibit different overall compositional differences compared to controls. There was a general trend toward the finding of increased abundances of bacteria involved in glutamate and GABA metabolism, and lower abundances of butyrate-producing bacteria in psychological disorders

## Introduction

Gut microbiome i.e symbiotic bacteria in the human gastrointestinal (GI) tract supply essential “metabolic machinery” for human bodies. Some define the gut microbiome as a ‘virtual organ’ [1] due to the extent to which it affects numerous facets of physiology through neurological, hormonal, and immunological pathways. The link between the intestinal microbiota and the central nervous system is referred to as the “bacteria-gut-brain axis” [2]. Even though they play a significant role in health maintenance, gut bacteria have been shown to disrupt homeostatic balance and influence the genesis and pathophysiology of a broad variety of diseases [3]. The enteric nervous system (ENS), also known as the “second brain,” is located in the body’s periphery and is responsible for facilitating the central nervous system’s (CNS) communication with the digestive tract [4].

“Anxiety disorders are the most frequent mental comorbidity in people with functional GI difficulties [5], and abdominal discomfort is one of the most known physical signs of sadness [9]. Successful top-down treatments using antidepressants and psychosocial therapies in the treatment of irritable bowel syndrome (IBS) [11] further define personality disorders as more than merely CNS disorders, but disorders with highly extensive systemic interconnections [6]. Psychiatry stands apart from other medical fields since there are no established biomarkers to aid in diagnosis or prognosis, and the root causes of most mental disorders are still a mystery. This indicates that the presentation of symptoms is the primary determinant of the classification of major mental diseases including mood and psychotic disorders [7].

There is growing evidence that the gut microbiota plays a role in the dysregulation of inflammation [8], oxidative stress [9], tryptophan metabolism [10], mitochondrial dysfunction [11], neurotransmitters [12], brain plasticity and neurotrophic factors [13], and metabolic processes [14]. Now that we know more about the functional potential of many bacteria [15], it is crucial to pinpoint the specific taxa that are disproportionately represented in patients with mental illnesses and that contribute to the dysregulation of the aforementioned systems. Changes in diet [16], antibiotics [17], probiotic supplements [18], and maybe even faecal microbial transplants [19]” are just few examples of how this information might inform the development of novel targeted treatment options. as well as insights into the etiology and the identification of clinically useful biomarkers. Now, there are a number of observational studies that have looked at whether or not the gut microbiota of persons with mental problems differs from that of healthy controls.

These data have been compiled in previous systematic studies for mood disorders [20], psychosis [21], and BD [22]. These showed that there was substantial variation in research methods and outcomes. There have been many new research published in this area since these reviews were written. This indicates the fast growth of research on gut microbiota and calls for a revised synthesis. To far, however, only a few studies have attempted to synthesize the information to determine if there are similarities or variances in gut microbiota composition across a variety of mental diseases. “Aetiology, potential diagnostic and prognostic biomarkers, and new treatment targets and strategies to alter the gut microbiota in these major personal disorders could be informed by a better understanding of which bacteria are differentially abundant across disorders, as well as those that may discriminate between disorders.

Therefore, we recently conducted a systematic review of the observational literature comparing the gut microbiota composition of persons with mental health difficulties with healthy controls.” The researchers wanted to see whether there was any consistency in the kind of changes seen in the makeup of different mental diseases, so they compared patients with mental illnesses to control volunteers for each ailment. Our secondary goal was to think about how these pathologic changes in composition may be functionally important in the context of these severe mental diseases.

## Methods

This review was written according to the guidelines established by Preferred Reporting Items for Systematic Reviews and Meta-Analyses (PRISMA). Not a single rule was broken, yet a more thorough search strategy did provide more relevant results. (figure1). Pubmed, Scopus, Embase, Web of Science, Ovid, Global Health, PsycINFO, etc. were searched thoroughly using the phrases “gut microbiota, psychological disorders, personality disorderscomposition, major depressive disorder, bipolar disorders, schizophrenia, etc.” Extensive searches were performed for all publications using inclusion and exclusion criteria.in the range of June 28th, 2018 and June 28th, 2023.

Before being tasked with the screening activity, reviewers received training in both full-text evaluation and assessment of simply the abstracts. The test was executed in an abstract manner using the Rayyan program. While one observer (AB) looked through all of the search results, three researchers (XX, HH, and JJ) independently reviewed 33.33 percent of the total hits twice. After reading the abstracts, the review committee got together to resolve their differences and create the final list of articles that needed to be evaluated in full. A full-text review was conducted using the Covidence program. Two independent reviewers, WW and YY, read the whole articles and rated them according to the criteria. When researchers weren’t sure whether or not a certain method was employed in an article, they went straight to the authors to ask for clarification. Members of the panel and reviewers from the scientific committee reached consensus on the final list of articles to be considered for review.

Included were original studies, literature reviews, scientific communications, systematic reviews, letters to the editor, and many additional preprints addressing the following areas related to gut microbiota and psychiatric disorders: (1) The role of the gut microbiome in depression (2) The role of the gut microbiome in bipolar disorder (3) Schizophrenia risk factors: the gut microbiome, (4) A link may be seen between the diversity of gut microbiota and personal disorders. Exclusion criteria were outlined as follows: Publications discovered in newspapers, magazines, blogs, and other non-academic venues; (1) written materials not in English; (2) documents related to issues not included in the inclusion requirements; (3) publications not written for an academic audience.

## Results

189 papers were discovered through a literature search using search criteria. There were 124 publications that were excluded because they were duplicates or similar. 65 different articles were first chosen. Following an examination of the titles and abstracts, thirty publications were removed. For 35 articles, full text management was done. Extra two papers were manually retrieved from references. There were 37 articles with full texts that could be read. 11 subpar articles were eliminated from the final evaluation. Finally, for this comprehensive assessment, 26 articles were chosen. (figure 1) (Table 1).

**Figure 1:**
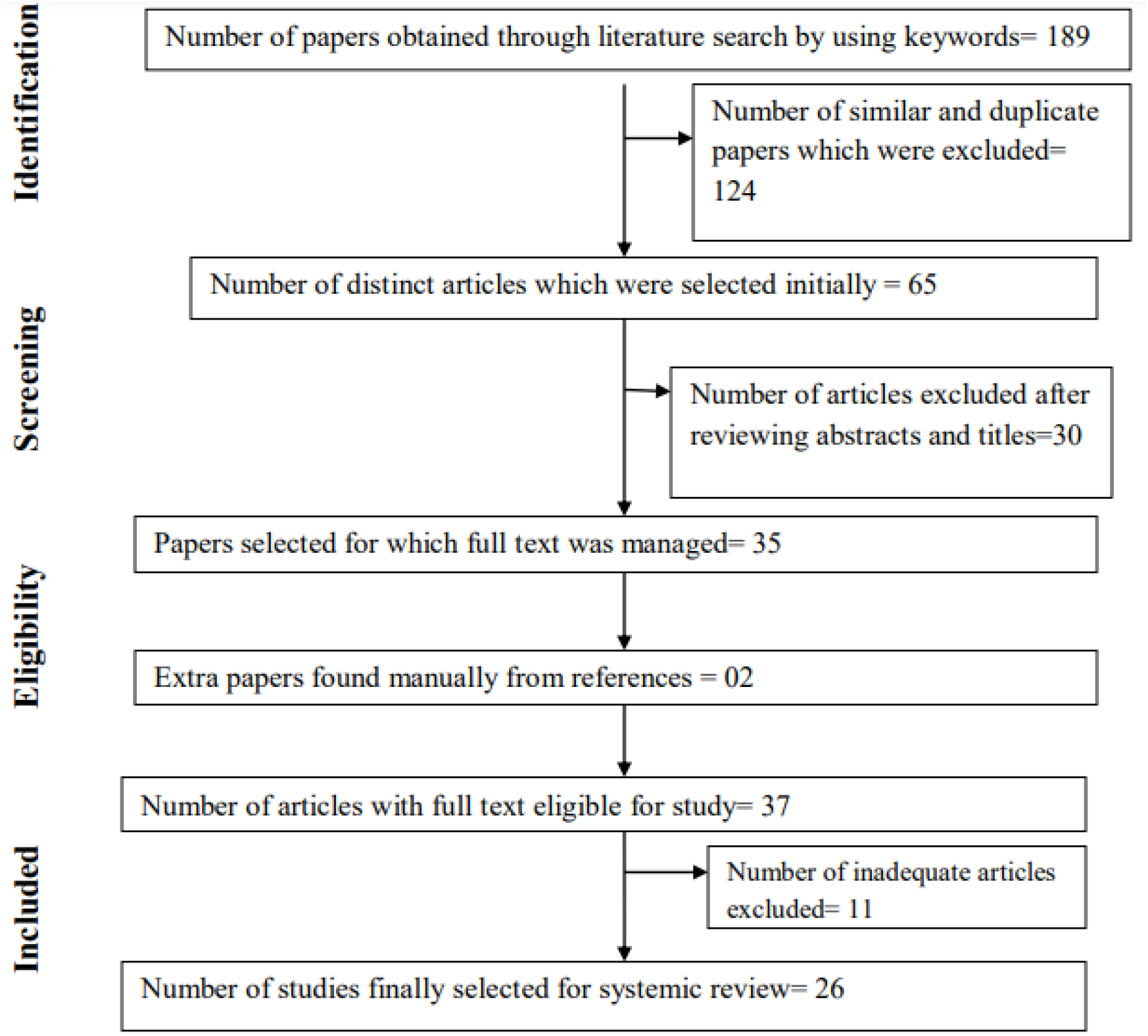
PRISMA flowchart for selection of studies in systematic review

**Table 1:** Important features of studies included in systematic review.

| S.N<br>o | Author<br>s with<br>year | Intension and<br>motive | Psychological<br>disorder<br>evaluated and<br>basic<br>methodology | Results | Discussion |
| --- | --- | --- | --- | --- | --- |
| 1 | Bai S<br>et al<br>2021 | This study set<br>out to test the<br>hypothesis that | MDD | Biomarkers for major<br>depressive disorder<br>(MDD) were discovered | These results<br>suggested that |
|  | [31] | inflammatory blood metabolites produced by the gut microbiota may act as biomarkers for clinical depression |  | using a technique that combines serum metabolomics with fecal microbial communities. Out of the 17 total genera identified, 14 were classified as Firmicutes. Analyses of gene expression differences revealed four significantly changed inflammatory-related pathways. | disruption of the phylum Firmicutes may play a role in the onset of depression by regulating the host's inflammatory response, and these five inflammation-related metabolites were identified as potential biomarkers that could be useful for future investigation into objective methods for diagnosing MDD. |
| 2 | Chen JJ et al, 2018 [32] | The purpose of this research was to learn | MDD | It was discovered that the gut microbiota composition changed with age, with the relative abundance of | If the study is fruitful, it will provide a new perspective to the quest for |
|  |  | more about the alterations in gut microbiota that occur in persons with MDD over time. |  | Firmicutes being significantly lower in young MDD patients compared to young healthy controls (HC) and the relative abundance of Bacteroidetes being significantly lower in middle-aged MDD patients compared to middle-aged HCs. | the root causes of MDD. |
| 3 | Chen YH et al, 2020 [33] | The purpose of this research was to define the gut microbiota and their functions in major depressive illness in females. | MDD | Bacteroidetes, proteobacteria, and Fusobacteria were overrepresented in the microbiomes of patients with MDD, whereas Firmicutes and Actinobacteria were consistently overrepresented in the microbiomes. | The etiology of major depressive illness in women may include microbiota and microbial metabolite alterations as possible microbiological targets and diagnostic indicators, respectively. |
| 4 | Chen JJ et al ,2018 [34] | The study's authors aimed to determine if there were | Female and male MDD patients were compared to healthy controls. | Each group was found to have 57 and 74 distinct operational taxonomic units, respectively. | These results demonstrated that persons with MDD had distinct gut flora based on their gender. |
|  |  | gender differences in the gut microbiome of people with MDD. |  | Female and male MDD patients had higher counts of Actinobacteria but lower counts of Bacteroidetes compared to healthy controls. The most notable differences in bacterial taxa between female and male MDD patients were seen in the phyla Actinobacteria and Bacteroidia. The major phylotypes of men and women with MDD were quite different. . | To what extent Actinobacteria and Bacteroidia may be used as sex-specific biomarkers for the diagnosis of MDD requires more study |
| 5. | Dong Z et al,2021 [35] | In order to establish a microbiome-based strategy for differential diagnosis,authors undertook a study to examine the variations in gut microbiota between typical instances of MDD and general anxiety disorders (GAD) | MDD, GAD | The microbiota richness and diversity of people with GAD were significantly different from those of those with HC. Sutterella and Fusicatenibacter were both substantially less common in MDD than in HC at the genus level, whereas the Christensenellaceae_R7_group and Fusicatenibacter were both much less common in GAD than in HC at the genus level. | GAD had a higher prevalence of Sutterella than MDD, but Faecalibacterium was much less common |
| 6. | Huang Y et al,2018 | Researchers studied the impact of | MDD | The MDD samples show the most dramatic | Depression may have a physiological |
|  | [36] | Firmicutes on people with MDD |  | reduction in the phylum Firmicutes. Thirteen taxonomic biomarkers from firmicutes have been shown to have a P value of less than 0.01. Depressed persons were shown to have significantly lower levels of Firmicutes and to vary from non-depressed people in 17 different KEGG pathways. | foundation in low-grade inflammation, and low levels of short-chain fatty acids may play a role in this which may have their origins in faulty Firmicutes. |
| 7. | Kelly JR e al,2016 [37] | Authors conducted the aforementioned research to determine whether or not changes in the gut microbiota's composition and function are at the root of the dysregulation of pathways.. | MDD | Depressive symptoms have been related to decreased diversity and richness of gut flora. Anxiety-like behaviors and alterations in tryptophan metabolism are seen when fecal microbiota from depressed people are transplanted into microbiota-depleted mice. all of which are hallmarks of depression in humans. | The results of this study point to the gut microbiota as a plausible target for the treatment and prevention of depressive symptoms |
| 8. | Lin P et al 2017 [38] | Authors looked at the possibility that the relative abundance of Prevotella and Klebsiella in | MDD | The diagnosis of MDD and other mental illnesses formerly depended on certain subjective survey methods. Extraction, PCR amplification, and | Prevotella and Klebsiella in the gut microbiome might serve as a diagnostic marker for |
|  |  | the gut microbiome might serve as a diagnostic marker for individuals with severe depressive disorder. |  | sequencing of fecal microbial communities from MDD patients on the Illumina Miseq platform revealed an increase in Firmicutes and a decrease in Bacteroidetes , as well as an increase in the numbers of bacteria belonging to the genera Prevotella, Klebsiella, Streptococcus, and Clostridium XI. Prevotella and Klebsiella percentage variations mirrored those seen on the Hamilton Depression Scale. | individuals with severe depressive disorder. |
| 9. | Liu RT et al, 2020 [39] | This study tried to found that among young individuals, depression was linked to a decline in anti-inflammatory gut flora. | The scientists compared the gut germs of 43 persons with MDD and 47 healthy controls, analyzing the gut microbiota of 90 young Americans in total | . The gut microbiota of those with MDD were different from those of healthy controls at many taxonomic levels. Class (Clostridia and Bacteroidia) and order (Clostridiales and Bacteroidales) levels of Firmicutes were reduced while Bacteroidetes were increased in people with MDD. | The results support the idea that a lack of butyrate-producing anti-inflammatory bacteria contributes to MDD, and they point to a connection between the gut microbiota and the chronic, low-grade<br>inflammation experienced by those with MDD. Similarly, the levels of Faecalibacterium and other members of the family Ruminococcaceae were lower in the MDD group compared to the healthy control group. |
| 10. | Liu Y et al, 2016 [40] | Clinical and pathophysiological features of irritable bowel syndrome (IBS), major depressive disorder (MDD), and IBS/MDD co-occurrence were analyzed in relation to fecal microbiota profiles | Immunohistochemical analyses of sigmoid tissue biopsy specimens to assess colonic mucosal inflammation | The etiology of both IBS-D and depression may be linked to changes in the fecal microbiota shared by patients with both disorders. There was a correlation between the degree of inflammation in the colon and the intensity of IBS symptoms. | The scientists identified three unique microbial profiles in humans, each of which has diagnostic potential for irritable bowel syndrome (IBS) or depression. |
| 11 | Mason BL et al,2020 [41] | To investigate the gut microbiota distributions of persons with co-occurring depression and anxiety, along with those with just depression or only anxiety, to see whether gut bacteria differently corresponds with unique clinical presentations. | MDD | Specifically, Bacteroides and the Clostridium leptum subgroup were found to be significantly different between clinical categories. But decreased Bacteroides may be more strongly linked to anxiety even in the absence of depression. | These variations point to the dispersion of gut microbiota as a potential source of insight into the etiology of co-occurring clinical manifestations. |
| 12 | Coello K et al, 2019 [42] | To analyse the gut microbiota diversity in freshly diagnosed BD individuals, those patients' unaffected immediate family members, and healthy people. | The gut microbiota composition was compared between 113 BD patients, 39 unaffected first-degree relatives, and 77 healthy persons by collecting stool samples from each group and profiling the microbiome using 16S rRNA gene amplicon sequencing. | The bacterial genus Flavonifractor has been linked by researchers to oxidative stress, inflammation, and host immune system dysfunction. to BD. | Our results may be impacted by residual confounding, and we cannot rule out the possibility that the higher smoking prevalence among individuals with BD is causal. |
| 13 | Evans SJ et al, 2017 [43] | Researchers looked into the gut microbiota of people with bipolar illness | BD | Better self-reported health outcomes on the Short Form Health Survey (SF12), the Patient Health | This is the first research to examine the connections between the |
|  |  | and compared them to those without the condition, as well as looked for correlations between the microbiome and disease load |  | Questionnaire (PHQ9), the Pittsburg Sleep Quality Index (PSQI), the Generalized Anxiety Disorder scale (GAD-7), and the Altman Mania Rating Scale (ASRM) were associated with higher fractional representation of Faecalibacterium among people with bipolar disorder, even after controlling for confounding variables.” | gut microbiota and a variety of mental conditions in a community of people with bipolar disorder. The findings lend credence to the theory that focusing on the microbiota might be a useful approach to treating bipolar illness |
| 14. | Hu S et al,2019 [44] | The researchers wanted to see if they could use changes in gut microbiota caused by quetiapine treatment as a biomarker for BD diagnosis and treatment success, and they also wanted to compare the microbiota of depressed patients with BD to that of healthy controls. | BD | Analysis of 16S ribosomal RNA gene sequences reveals that BD patients' and HCs' gut microbiomes differ in terms of both composition and diversity. The phylum Bacteroidetes predominates in the bacterial communities of patients with BD, whereas the phylum Firmicutes predominates in those with HCs. Butyrate-producing bacteria are present in lower numbers in untreated patients. The microbiota composition of persons with BD is changed by quetiapine treatment. | The findings provide new light on the role of the gut microbiota in BD and are the first to evaluate microbial changes after quetiapine therapy. Biomarkers derived from the gut microbiota's makeup may one day help in the diagnosis of BD and the estimation of therapeutic efficacy, however further study is required to prove this. |
| 15 | Lai WT et al 2021 [45] | The goal of the study by ABC et al. was to see whether changes in the gut microbiota in people with bipolar illness who are experiencing a severe depressive episode right now might be uncovered by using shotgun metagenomics | BD | The microbial tryptophan biosynthesis and metabolism pathway (MiTBamp) is of particular interest because to the links it has been demonstrated to have between BPD, the microbiome, the gut, and the brain. The feces of 25 people with BPD and 28 healthy controls (HCs) were sequenced using shotgun metagenomics (SMS). Using the Random Forests (RF) and Boruta algorithm, authors developed a classification model to better understand the alterations in the GI microbiome and MiTBamp in BPD | Changes in the microbiome have been suggested as a biomarker that might be used to distinguish BPD sufferers from HCs. |
| 16 | McIntyre RS et al, 2021 [46] | To better understand the gut microbiome of persons with bipolar illness | Subjects (aged 18-65) who met DSM-5 criteria for BD and healthy controls (HC) of the same age and sex but without a history of mental or serious medical illnesses both provided fecal samples. We employed microbial community analysis (ANCOM) and cluster analysis to look at how different types of microorganisms in the gut are related to each other, as well as how food affected the microbiome overall | Participants with BD had a higher frequency of Clostridiaceae OTUs compared to HC, while participants with BD-II had a higher prevalence of Collinsella OTUs compared to BD-I subjects. Cluster analysis showed that neither the diagnosis nor the diet had any significant impact on the gut microbiota's overall makeup. | According to the results of this research, the gut microbiota of people with BD may be different from that of those without the condition, namely due to a higher abundance of Clostridiaceae and Collinsella |
| 17 | Rong H et al,2019 [47] | Researchers conducted a study on the gut microbiota of persons with major depressive disorder, bipolar disorder who are now experiencing a major depressive episode, and healthy controls. | BD | The phyla Firmicutes and Actinobacteria had substantial increases in abundance in the MDD and BPD groups, whereas the phylum Bacteroidetes saw a major decrease | We postulate that the gut microbiota contributes to the etiology of MDD and BPD and that certain bacterial variants may serve as biomarkers for these conditions based on the findings of our study |
| 18 | Zheng P et al,2020 [48] | Researchers conducted a study on the markers of the gut microbiota to distinguish unipolar from bipolar disorder | BD | The microbiomes of the three groups were found to be qualitatively different. Changes in covarying OTUs of the Bacteroidaceae family are seen in MDD when compared to HCs, while changes in the Lachnospiraceae, Prevotellaceae, and Ruminococcaceae families are observed in BD. A signature of 26 OTUs was demonstrated to be able to differentiate between patients with | Collectively, we discover that MDD is distinguished from BD and HCs by its distinctive gut microbial composition, and we provide a novel marker panel for distinguishing MDD from BD based on patterns in the |
|  |  |  |  | MDD and those with BD or HCs, with AUC values of 0.961–0.986 in discovery sets and 0.702–0.741 in validation sets. The severity of the disease is also connected with 4 of the 26 microbiological indicators in MDD and BD | gut microbiome |
| 19 | Li S et al, 2020 [49] | Scientists investigated whether or not there was a link between alterations in gut flora and the manifestation of schizophrenia | Authors evaluated the associations between changed gut microbiota and the severity of symptoms in 82 SZ patients by comparing their fecal microbiota with that of 80 demographically comparable normal controls (NCs) using 16S rRNA sequencing | Results showed that the SZ group had a different microbiota in their digestive tract. | It showed, for the first time, that *Succinivibrio* and *Corynebacterium* were linked to more severe symptoms, suggesting that these bacteria might serve as useful novel biomarkers in the diagnosis of SZ. |
| 20 | Li S et al, 2021 [50] | Researchers set out to learn more about how the composition of one's gut microbiota | SCZ | They found that the abundance of *Veillonella* was much higher in SZ patients compared to NCs, but the abundance of *Ruminococcus* and *Roseburia* was much | Results suggest that the gut bacteria may play a role in SZ through influencing |
|  |  | might affect cognitive performance |  | lower in SZ patients. Low-frequency fluctuation amplitude was also considerably bigger in SZ patients compared to NCs, despite decreased gray matter volume (GMV) and regional homogeneity (ReHo). | brain anatomy and function. This research sheds light on the neuropathology that underlies SZ. |
| 21 | Ma X et al,2020 [51] | The pathophysiology of schizophrenia (SCZ) was investigated by authors who looked into possible links between the gut microbiota and SCZ | SCZ | Significant changes were seen between FSCZ and TSCZ patients and HCs for the families Christensenellaceae, Enterobacteriaceae, Pasteurellaceae, and Turicibacteraceae, and for the genus Escherichia. Significant changes in the makeup of gut microbes (such as Enterococcaceae and Lactobacillaceae) were also discovered between TSCZ and FSCZ patients. Intriguingly, we discovered that the abnormal volume of the right middle frontal gyrus (rMFG) in SCZ patients was coupled with particular SCZ-associated microbiota | Results build on previous research and provide more evidence that the gut microbiome may play a role in the pathogenesis of SCZ by influencing brain anatomy |
| 22 | Manchia M et al,2021 [52] | Authors looked at the connection between changes in the gut microbiota and the emergence of antipsychotic drug resistance | Determine whether there were any differences in the gut microbiota composition of SCZ patients who responded differently to antipsychotics (18 | findings showed that SCZ and HC had distinct gut microbiota. with some taxonomic levels of bacteria being exclusive to one group or the other. Results were similar regardless of the kind of antipsychotic used or how the patient | Results raise the possibility that the composition of gut microbiota serves as a biosignature for both SCZ and TRS |
|  |  | in patients with schizophrenia. | patients with treatment-resistant schizophrenia (TRS) and 20 patients who responded to antipsychotics). | responded to therapy. |  |
| 23 | Miao Y et al, 2021 [53] | Authors . examined the relationship between gut microbiota and blood folic acid level in first-episode, drug-free individuals with schizophrenia (SCZ). | SCZ | The general psychopathology score was positively correlated with the interaction term between folic acid and genus Bifidobacterium in SCZ patients, but the interaction term was not correlated with cognitive function (both $P > 0.05$). | Psychiatric symptoms in first-episode, antibiotic-naïve patients were linked to low blood folic acid levels and a high abundance of Bifidobacterium. drug-free SCZ patients. This suggests that these substances may serve as objective indicators of psychiatric symptoms. |
| 24 | Nguyen TT et al, 2019 [54] | To compare people with chronic schizophrenia and non-psychiatric controls (NCs) with similar demographics | In this research, 50 people were analyzed, including 25 people with chronic schizophrenia and 25 non-psychiatric controls (NCs) with similar demographics | Proteobacteria were discovered to be much lower in the brains of people with schizophrenia compared to those of healthy controls. Schizophrenia was associated with an increase in the species Anaerococcus and a reduction in the genera Haemophilus, Sutterella, and Clostridium. | Findings of study lend credence to the idea that patients with chronic schizophrenia have a unique gut microbiota compared to the general population and call attention |
|  |  |  |  | Ruminococcaceae abundance was linked with reduced negative symptom severity in people with schizophrenia; More Bacteroides was linked to more severe depression, whereas more Coprococcus meant a higher chance of heart disease. | to the need for more large-scale longitudinal studies of the microbiome and schizophrenia |
| 25 | Nguyen TT et al,2021 [55] | Using 16S rRNA sequencing, authors examined the gut microbiota composition and functional capacity in 48 people with chronic schizophrenia and 48 healthy controls (NCs) who were similar to the patients in terms of sequencing plate, age, sex, body mass index (BMI), and antibiotic usage. | In order to find differentially abundant microorganisms and pathways, authors used a novel compositionally-aware approach that included reference frames | Results suggested that Lachnospiraceae is linked to schizophrenia. Both the trimethylamine-N-oxide reductase and Kdo2-lipid A biosynthesis functional pathways were shown to be altered in people with schizophrenia. Pro-inflammatory cytokines and an elevated risk of cardiovascular disease were both connected to these metabolic pathways in patients with schizophrenia | These findings stimulate new lines of inquiry, such as whether the microbiota influence the pathophysiology of the disease by influencing immune signaling/response and lipid/glucose regulation functioning pathways. |
| 26. | Pan R et al,2020 [56] | In order to investigate the potential of the gut microbiome as a non-invasive biomarker for schizophrenia | SCZ | These findings demonstrate differences between SCZ patients and healthy controls, and between patients in the acute phase and those in remission, highlighting the potential of the gut | SCZ-specific gut microbiota characteristics provide insights into illness prognosis and enable more |
|  |  | (SCZ), authors undertook a research comparing the gut microbiota features of individuals with acute and remission SCZ |  | microbiota as a non-invasive diagnostic tool. | precise treatment |

### Gut microbiota and Major Depressive Disorder

Biomarkers for major depressive disorder (MDD) were discovered using a technique that combines serum metabolomics with fecal microbial communities. Out of the 17 total genera identified, 14 were classified as Firmicutes. Analyses of gene expression differences revealed four significantly changed inflammatory-related pathways. These results suggested that disruption of the phylum Firmicutes may play a role in the onset of depression by regulating the host’s inflammatory response, and these five inflammation-related metabolites were identified as potential biomarkers that could be useful for future investigation into objective methods for diagnosing MDD. [31]

It was discovered that the gut microbiota composition changed with age, with the relative abundance of Firmicutes being significantly lower in young MDD patients compared to young HCs and the relative abundance of Bacteroidetes being significantly lower in middle-aged MDD patients compared to middle-aged HCs. If our study is fruitful, it will provide a new perspective to the quest for the root causes of MDD. [32] Bacteroidetes, proteobacteria, and Fusobacteria were overrepresented in the microbiomes of patients with MDD, whereas Firmicutes and Actinobacteria were consistently overrepresented in the microbiomes. The etiology of major depressive illness in women may include microbiota and microbial metabolite alterations as possible microbiological targets and diagnostic indicators, respectively. [33]

The study’s authors aimed to determine if there were gender differences in the gut microbiome of people with MDD. Female and male MDD patients were compared to healthy controls, and each group was found to have 57 and 74 distinct operational taxonomic units, respectively. Female and male MDD patients had higher counts of Actinobacteria but lower counts of Bacteroidetes compared to healthy controls. The most notable differences in bacterial taxa between female and male MDD patients were seen in the phyla Actinobacteria and Bacteroidia. The major phylotypes of men and women with MDD were quite different. These results demonstrated that persons with MDD had distinct gut flora based on their gender. To what extent Actinobacteria and Bacteroidia may be used as sex-specific biomarkers for the diagnosis of MDD requires more study.[34]

In order to establish a microbiome-based strategy for differential diagnosis, Dong Z et al,2021 et al. undertook a study to examine the variations in gut microbiota between typical instances of various disorders. The microbiota richness and diversity of people with GAD were significantly different from those of those with HC. Sutterella and Fusicatenibacter were both substantially less common in MDD than in HC at the genus level, whereas the Christensenellaceae_R7_group and Fusicatenibacter were both much less common in GAD than in HC at the genus level. GAD had a higher prevalence of Sutterella than MDD, but Faecalibacterium was much less common. Differential diagnosis and targeted treatment for MDD and GAD may be aided by the distinct gut-microbiome profile identified in this research.[35]

Huang Y et al,2018 studied the impact of Firmicutes on people with MDD. Patients with MDD were shown to have decreased alpha diversity indices compared to healthy controls. The MDD samples show the most dramatic reduction in the phylum Firmicutes. Thirteen taxonomic biomarkers from firmicutes have been shown to have a P value of less than 0.01. Depressed persons were shown to have significantly lower levels of Firmicutes and to vary from non-depressed people in 17 different KEGG pathways. Depression may have a physiological foundation in low-grade inflammation, and low levels of short-chain fatty acids may play a role in this. which may have their origins in faulty Firmicutes.[36]

Kelly JR e al,2016 conducted the aforementioned research to determine whether or not changes in the gut microbiota’s composition and function are at the root of the dysregulation of these pathways. Depressive symptoms have been related to decreased diversity and richness of gut flora. Anxiety-like behaviors and alterations in tryptophan metabolism are seen when fecal microbiota from depressed people are transplanted into microbiota-depleted mice. all of which are hallmarks of depression in humans. The results of these studies point to the gut microbiota as a plausible target for the treatment and prevention of depressive symptoms. [37]

Researchers looked at the possibility that the relative abundance of Prevotella and Klebsiella in the gut microbiome might serve as a diagnostic marker for individuals with severe depressive disorder. The diagnosis of MDD and other mental illnesses formerly depended on certain subjective survey methods. Extraction, PCR amplification, and sequencing of fecal microbial communities from MDD patients on the Illumina Miseq platform revealed an increase in Firmicutes and a decrease in Bacteroidetes, as well as an increase in the numbers of bacteria belonging to the genera Prevotella, Klebsiella, Streptococcus, and Clostridium XI.Prevotella and Klebsiella percentage variations mirrored those seen on the Hamilton Depression Scale. [38]

A study found that among young individuals, depression was linked to a decline in anti-inflammatory gut flora. The scientists compared the gut germs of 43 persons with MDD and 47 healthy controls, analyzing the gut microbiota of 90 young Americans in total. The gut microbiota of those with MDD were different from those of healthy controls at many taxonomic levels. Class (Clostridia and Bacteroidia) and order (Clostridiales and Bacteroidales) levels of Firmicutes were reduced while Bacteroidetes were increased in people with MDD. The results support the idea that a lack of butyrate-producing anti-inflammatory bacteria contributes to MDD, and they point to a connection between the gut microbiota and the chronic, low-grade inflammation experienced by those with MDD.Similarly, the levels of Faecalibacterium and other members of the family Ruminococcaceae were lower in the MDD group compared to the healthy control group. [39]

Clinical and pathophysiological features of irritable bowel syndrome (IBS), major depressive disorder (MDD), and IBS/MDD co-occurrence were analyzed in relation to fecal microbiota profiles using immunohistochemical analyses of sigmoid tissue biopsy specimens to assess colonic mucosal inflammation. The etiology of both IBS-D and depression may be linked to changes in the fecal microbiota shared by patients with both disorders. There was a correlation between the degree of inflammation in the colon and the intensity of IBS symptoms. The scientists identified three unique microbial profiles in humans, each of which has diagnostic potential for irritable bowel syndrome (IBS) or depression.[40]

Mason BL et al,2020 investigated the gut microbiota distributions of persons with co-occurring depression and anxiety, along with those with just depression or only anxiety, to see whether gut bacteria differently corresponds with unique clinical presentations. Specifically, Bacteroides and the Clostridium leptum subgroup were found to be significantly different between clinical categories. But decreased Bacteroides may be more strongly linked to anxiety even in the absence of depression. These variations point to the dispersion of gut microbiota as a potential source of insight into the etiology of co-occurring clinical manifestations. [41].

### Gut microbiota in Bipolar Disorder

The gut microbiota composition was compared between 113 BD patients, 39 unaffected first-degree relatives, and 77 healthy persons by collecting stool samples from each group and profiling the microbiome using 16S rRNA gene amplicon sequencing. The bacterial genus Flavonifractor has been linked by researchers to oxidative stress, inflammation, and host immune system dysfunction. to BD. Our results may be impacted by residual confounding, and we cannot rule out the possibility that the higher smoking prevalence among individuals with BD is causal.[42]

Researchers looked into the gut microbiota of people with bipolar illness and compared them to those without the condition, as well as looked for correlations between the microbiome and disease load. “Better self-reported health outcomes on the Short Form Health Survey (SF12), the Patient Health Questionnaire (PHQ9), the Pittsburg Sleep Quality Index (PSQI), the Generalized Anxiety Disorder scale (GAD-7), and the Altman Mania Rating Scale (ASRM) were associated with higher fractional representation of Faecalibacterium among people with bipolar disorder, even after controlling for confounding variables.” This is the first research to examine the connections between the gut microbiota and a variety of mental conditions in a community of people with bipolar disorder. The findings lend credence to the theory that focusing on the microbiota might be a useful approach to treating bipolar illness.[43]

The researchers wanted to see if they could use changes in gut microbiota caused by quetiapine treatment as a biomarker for BD diagnosis and treatment success, and they also wanted to compare the microbiota of depressed patients with BD to that of healthy controls. Analysis of 16S ribosomal RNA gene sequences reveals that BD patients’ and HCs’ gut microbiomes differ in terms of both composition and diversity. The phylum Bacteroidetes predominates in the bacterial communities of patients with BD, whereas the phylum Firmicutes predominates in those with HCs. Butyrate-producing bacteria are present in lower numbers in untreated patients. The microbiota composition of persons with BD is changed by quetiapine treatment. The findings provide new light on the role of the gut microbiota in BD and are the first to evaluate microbial changes after quetiapine therapy. Biomarkers derived from the gut microbiota’s makeup may one day help in the diagnosis of BD and the estimation of therapeutic efficacy, however further study is required to prove this.[44]

The goal of the study by Lai WT et al 2021. was to see whether changes in the gut microbiota in people with bipolar illness who are experiencing a severe depressive episode right now might be uncovered by using shotgun metagenomics. The microbial tryptophan biosynthesis and metabolism pathway (MiTBamp) is of particular interest because to the links it has been demonstrated to have between BPD, the microbiome, the gut, and the brain. The feces of 25 people with BPD and 28 healthy controls (HCs) were sequenced using shotgun metagenomics (SMS). Using the Random Forests (RF) and Boruta algorithm, we developed a classification model to better understand the alterations in the GI microbiome and MiTBamp in BPD. Changes in the microbiome have been suggested as a biomarker that might be used to distinguish BPD sufferers from HCs.[45]

To better understand the gut microbiome of persons with bipolar illness subjects (aged 18-65) who met DSM-5 criteria for BD and healthy controls (HC) of the same age and sex but without a history of mental or serious medical illnesses both provided fecal samples. We employed microbial community analysis (ANCOM) and cluster analysis to look at how different types of microorganisms in the gut are related to each other, as well as how food affected the microbiome overall. participants with BD had a higher frequency of Clostridiaceae OTUs compared to HC, while participants with BD-II had a higher prevalence of Collinsella OTUs compared to BD-I subjects. Cluster analysis showed that neither the diagnosis nor the diet had any significant impact on the gut microbiota’s overall makeup. According to the results of this research, the gut microbiota of people with BD may be different from that of those without the condition, namely due to a higher abundance of Clostridiaceae and Collinsella.[46]

Rong H et al,2019 conducted a study on the gut microbiota of persons with major depressive disorder, bipolar disorder who are now experiencing a major depressive episode, and healthy controls. The phyla Firmicutes and Actinobacteria had substantial increases in abundance in the MDD and BPD groups, whereas the phylum Bacteroidetes saw a major decrease. We postulate that the gut microbiota contributes to the etiology of MDD and BPD and that certain bacterial variants may serve as biomarkers for these conditions based on the findings of our study. [47]

Zheng P et al,2020 conducted a study on the markers of the gut microbiota to distinguish unipolar from bipolar disorder. The microbiomes of the three groups were found to be qualitatively different. Changes in covarying OTUs of the Bacteroidaceae family are seen in MDD when compared to HCs, while changes in the Lachnospiraceae, Prevotellaceae, and Ruminococcaceae families are observed in BD. A signature of 26 OTUs was demonstrated to be able to differentiate between patients with MDD and those with BD or HCs, with AUC values of 0.961–0.986 in discovery sets and 0.702–0.741 in validation sets. The severity of the disease is also connected with 4 of the 26 microbiological indicators in MDD and BD. Collectively, we discover that MDD is distinguished from BD and HCs by its distinctive gut microbial composition, and we provide a novel marker panel for distinguishing MDD from BD based on patterns in the gut microbiome.[48]

### Gut microbiota in Schizophrenia

Scientists investigated whether or not there was a link between alterations in gut flora and the manifestation of schizophrenia. They evaluated the associations between changed gut microbiota and the severity of symptoms in 82 SZ patients by comparing their fecal microbiota with that of 80 demographically comparable normal controls (NCs) using 16S rRNA sequencing. Their results showed that the SZ group had a different microbiota in their digestive tract. We also showed, for the first time, that Succinvibrio and Corynebacterium were linked to more severe symptoms, suggesting that these bacteria might serve as useful novel biomarkers in the diagnosis of SZ.[49]

Li S et al, 2021 set out to learn more about how the composition of one’s gut microbiota might affect cognitive performance. We found that the abundance of Veillonella was much higher in SZ patients compared to NCs, but the abundance of Ruminococcus and Roseburia was much lower in SZ patients. Low-frequency fluctuation amplitude was also considerably bigger in SZ patients compared to NCs, despite decreased gray matter volume (GMV) and regional homogeneity (ReHo). Our results suggest that the gut bacteria may play a role in SZ through influencing brain anatomy and function. This research sheds light on the neuropathology that underlies SZ.[50]

The pathophysiology of schizophrenia (SCZ) was investigated by Ma X et al,2020, who looked into possible links between the gut microbiota and SCZ. Significant changes were seen between FSCZ and TSCZ patients and HCs for the families Christensenellaceae, Enterobacteriaceae, Pasteurellaceae, and Turicibacteraceae, and for the genus Escherichia. Significant changes in the makeup of gut microbes (such as Enterococcaceae and Lactobacillaceae) were also discovered between TSCZ and FSCZ patients. Intriguingly, we discovered that the abnormal volume of the right middle frontal gyrus (rMFG) in SCZ patients was coupled with particular SCZ-associated microbiota. Our results build on previous research and provide more evidence that the gut microbiome may play a role in the pathogenesis of SCZ by influencing brain anatomy.[51]

In a recent research, Manchia M et al,2021 looked at the connection between changes in the gut microbiota and the emergence of antipsychotic drug resistance in patients with schizophrenia. The aim of this study was to determine whether there were any differences in the gut microbiota composition of SCZ patients who responded differently to antipsychotics (18 patients with treatment-resistant schizophrenia (TRS) and 20 patients who responded to antipsychotics). Our findings showed that SCZ and HC had distinct gut microbiota. with some taxonomic levels of bacteria being exclusive to one group or the other. Results were similar regardless of the kind of antipsychotic used or how the patient responded to therapy. Our findings raise the possibility that the make-up of gut microbiota serves as a biosignature for both SCZ and TRS. [52]

In a study Miao Y et al,2021 examined the relationship between gut microbiota and blood folicacid level in first-episode, drug-free individuals with schizophrenia (SCZ). The general psychopathology score was positively correlated with the interaction term between folic acid and genus Bifidobacterium in SCZ patients, but the interaction term was not correlated with cognitive function (both P>0.05). Psychiatric symptoms in first-episode, antibiotic-naive patients were linked to low blood folic acid levels and a high abundance of Bifidobacterium. drug-free SCZ patients. This suggests that these substances may serve as objective indicators of psychiatric symptoms.[53]

In a research, 50 people were analyzed, including 25 people with chronic schizophrenia and 25 non-psychiatric controls (NCs) with similar demographics. Proteobacteria were discovered to be much lower in the brains of people with schizophrenia compared to those of healthy controls. Schizophrenia was associated with an increase in the species Anaerococcus and a reduction in the genera Haemophilus, Sutterella, and Clostridium. Ruminococcaceae abundance was linked with reduced negative symptom severity in people with schizophrenia; More Bacteroides was linked to more severe depression, whereas more Coprococcus meant a higher chance of heart disease. Our findings lend credence to the idea that patients with chronic schizophrenia have a unique gut microbiota compared to the general population and call attention to the need for more large-scale longitudinal studies of the microbiome and schizophrenia.[54]

Using 16S rRNA sequencing, Nguyen TT et al,2021 examined the gut microbiota composition and functional capacity in 48 people with chronic schizophrenia and 48 healthy controls (NCs) who were similar to the patients in terms of sequencing plate, age, sex, body mass index (BMI), and antibiotic usage. In order to find differentially abundant microorganisms and pathways, we used a novel compositionally-aware approach that included reference frames, and our results suggested that Lachnospiraceae is linked to schizophrenia. Both the trimethylamine-N-oxide reductase and Kdo2-lipid A biosynthesis functional pathways were shown to be altered in people with schizophrenia. Pro-inflammatory cytokines and an elevated risk of cardiovascular disease were both connected to these metabolic pathways in patients with schizophrenia. These findings stimulate new lines of inquiry, such as whether the microbiota influence the pathophysiology of the disease by influencing immune signaling/response and lipid/glucose regulation functioning pathways.[55]

In order to investigate the potential of the gut microbiome as a non-invasive biomarker for schizophrenia (SCZ), Pan R et al,2020 undertook a research comparing the gut microbiota features of individuals with acute and remission SCZ. These findings demonstrate differences between SCZ patients and healthy controls, and between patients in the acute phase and those in remission, highlighting the potential of the gut microbiota as a non-invasive diagnostic tool. In addition, SCZ-specific gut microbiota characteristics provide insights into illness prognosis and enable more precise treatment.[56]

## Discussion

As far as we are aware, this is the first systematic review of the research on the issue of gut microbiota composition and major personal disorders such as MDD, BD, and SZ. We did discover widespread differences in the gut microbiota makeup of patients and controls under each category of personal disorder. We also found that there are distinct bacterial taxa that had differing abundances in patients with these three psychiatric illnesses compared to healthy controls. We found a great deal of variation in study designs and reporting, such as in the inclusion and exclusion of study populations, sampling feces for study of gut microbiota; taking into account or adjusting for important factors known to impact gut microbiota composition; storing feces; processing feces; analyzing feces. Last but not least, we conducted a quality assessment of the included research; the results lend credence to the creation of norms for the execution and reporting of microbiome-related studies.

Our analysis showed that patients with all three mental disorders had higher levels of Eggerthella and Lactobacillus and lower levels of Coprococcus compared to controls. Multiple diseases also shared several bacterial genera. Additionally, MDD and SZ were shown to have an increase in Escherichia coli, Shigella, and Veillonella whereas SZ and BD shared an increase in Megasphaera and a reduction in Roseburia. Enterococcus faecium,Flavonifractor, and Streptococcus were associated with MDD and BD, whereas decreased Faecalibacterium and Ruminococcus were associated with BD.

While there were certain taxa that were differently abundant between patients and controls for all three mental illnesses, there were also some taxa that were differentially abundant just for one condition. Patients with MDD typically had higher levels of Alistipes and Parabacteroides and lower levels of Prevotella, whereas patients with BD typically had higher levels of Bifidobacterium and Oscillibacter, and patients with SZ typically had higher levels of Prevotella and lower levels of Bacteroides, Haemophilus, and Streptococcus.

Our analysis showed an increase in lactic acid-producing bacteria in all three microbiome types (MDD, BD, and SZ). Cases of the three most common mental illnesses had a greater prevalence of the genus Lactobacillus. Other lactic acid manufacturers, such as Enterococcus and Streptococcus, were also observed to be more abundant in patients with MDD, BD, SZ, and Bifidobacterium, respectively. Metabolic regulation, pathogen protection, and immunomodulatory actions are only some of the host-beneficial effects attributed to these bacteria [24-28]. Cross-feeding occurs when bacteria that make lactate also offer it to bacteria that utilize lactate as a substrate to produce metabolites such the SCFA butyrate [29]. However, there are situations when host health is compromised by lactate generation and use. Lactic acid buildup in the intestines has been linked to acidosis, cardiac arrhythmias, and neurotoxicity [29, 30]. Improving results in moderate to severe MDD [32] also raise butyrate-producing bacteria [33], suggesting that mitochondrial dysfunction may be at the root of many mental diseases. There is proof that enriched bacteria may affect GABA metabolism [32,33]. Our analysis also revealed that all three mental diseases shared an increase in the abundance of bacteria involved in the metabolism of glutamate and -aminobutyric acid (GABA). Again, there was less evidence to imply that this pattern was linked to any one ailment in particular; rather, elevated Lactobacillus was a trait shared by a wide range of conditions. MDD was characterized by increased abundances of bacteria involved in glutamate and GABA metabolism, including Alistipes and Parabacteroides, whereas BD was characterized by increased abundances of Bifidobacterium and Enterococcus and both MDD and BD were characterized by increased abundances of Bacteroides and Streptococcus. Genes encoding glutamate decarboxylase (GAD) enzymes, which catalyze the conversion of L-glutamate to GABA, are found in the lactic acid bacteria Lactobacillus, Bifidobacterium, and Enterococcus [33, 34]. Despite getting less attention from researchers, it seems that Eggerthella may alter glutamate metabolism through GAD, since elevated levels of this bacteria have been linked to abnormalities in glutamate metabolism in children with autism spectrum disorder [35]. GABA production has also been associated with the bacterial species Bacteroides, Escherichia, and Parabacteroides [36]. Increased numbers of these gut bacteria have been linked to an array of mental health conditions, suggesting that they may promote glutamate depletion and GABA production. various bacterial abundances may have various pathophysiological effects, although this has yet to be proved. The complexity of the role played by the human gut microbiota in health and disease necessitates the use of multi-omics methods. More study is needed to determine whether or not alterations in the gut microbiota are primarily driven by pathophysiology or by common risk factors like nutrition, or whether or not they are driven by both. Changes in gut microbiota and their possible influence on disease development may be recorded in future longitudinal cohort studies. The biochemical and molecular impact that certain bacterial taxa have on host health and sickness may be elucidated via intervention studies. There is a significant lack of repeatability and heterogeneity in study methodologies. This literature review sheds insight on the wide discrepancy between how human microbiome data is collected and reported.

Consensus on best-practice methods is always shifting or being updated, which makes establishing or identifying ‘gold-standards’ difficult due to the quick rate at which this sector is expanding. Limited resources often force researchers to use suboptimal research designs and settle for limited sample sizes. Since there are no well-established methods for calculating power, it is not always clear whether microbiome research have adequate power to identify differences. There is an urgent need for clarity in reporting of microbiome research and for the consideration of these restrictions within individual investigations because of the impact of different microbiome-related study procedures on study outcomes [37-39]. Mutations in the gut microbiota have been linked to a wide range of factors, not all of which are within the control of the person. It is crucial to collect information on these factors in order to assess and interpret findings correctly. The fact that gut microbiota composition is often a secondary research outcome may account for the lack of collection and consideration of factors in study design. Possibly improving methodological consistency and repeatability [40-43] is the ‘Strengthening the Organizing and Reporting of Microbiome studies’ (STORMS) tool [44-52], a newly created checklist for the reporting of human microbiome research. [57,58].

### Limitations and strengths

Several aspects of our present review’s methodology affect how these findings should be interpreted. One problem is that the included studies were cross-sectional, thus no causation can be inferred and no time-dependent changes in the gut microbiota can be accounted for. Second, when broken down by country of origin, Chinese studies are noticeably overrepresented. This imbalance in sample area may have affected the findings of our synthesis since various geographical locations are linked to diverse microbial compositions. This study focused only on characterizing gut bacteria, despite mounting evidence that these organisms have an impact on host psychology and behavior. The GI tract is home to a wide variety of microorganisms, including archaea, viruses, bacteriophages, and fungus; research into the effect these microbes may have on host mental health is only beginning. Fourth, it is well acknowledged that using just compositional data in studies of the gut microbiome (mostly using 16S) has significant drawbacks, including decreased sensitivity and resolution. Despite these caveats, the information presented here lays a crucial groundwork for future research into the role of the gut microbiota in psychiatry. As additional studies are conducted and published utilizing omics methodologies (such as metagenomics, metabolomics, and meta-transcriptomics), we will be able to get a deeper understanding of the gut microbiome beyond what is now known about its makeup. Psychiatry is currently lacking in biomarkers for diagnosis and prognosis as well as a strong understanding of the aetiology of disease. may gain greatly from this. Fifth, there is the possibility of unmeasured confounding skewing our syntheses. Few research even attempted to account for possible confounding by collecting covariate data consistently across trials. In conclusion, our synthesis highlighted a number of methodological differences across research. Although it is beyond the scope of this analysis to go into detail about these distinctions, we have included a summary of the most important ones in the appendices.

## Conclusion

Our systematic review did find that psychological disorders appeared to exhibit different overall compositional differences compared to controls. There was a general trend toward the finding of increased abundances of bacteria involved in glutamate and GABA metabolism, and lower abundances of butyrate-producing bacteria in psychological disorders. Future research using multi-omics methodologies is required to elucidate the implications of compositional and taxonomic changes for mental disorder pathophysiology and etiology. Our results suggest that certain bacterial genera may one day be useful for diagnosis and prognosis; additional study is needed to validate this. Furthermore, these results may lend credence to alternative therapeutic approaches, include diet plans that aim to correct an imbalance in the gut microbiome. The study of the human microbiome has a clear and urgent need for standardized reporting and techniques.

## Conflict of interest

The authors declare no conflict of interest

## Data Availability

All data produced in the present work are contained in the manuscript

## Acknowledgements

We extend our gratitude to President Prof. Ismail Matalka and Dean Prof. K. Bairy for their invaluable assistance and support throughout this research.

## Notes

### Competing Interest Statement

The authors have declared no competing interest.

### Funding Statement

This study did not receive any funding

